# Prediction of COVID-19 Spreading Profiles in South Korea, Italy and Iran by Data-Driven Coding

**DOI:** 10.1101/2020.03.08.20032847

**Authors:** Choujun Zhan, Chi K. Tse, Zhikang Lai, Tianyong Hao, Jingjing Su

## Abstract

This work applies a data-driven coding method for prediction of the COVID-19 spreading profile in any given population that shows an initial phase of epidemic progression. Based on the historical data collected for COVID-19 spreading in 367 cities in China and the set of parameters of the augmented Susceptible-Exposed-Infected-Removed (SEIR) model obtained for each city, a set of profile codes representing a variety of transmission mechanisms and contact topologies is formed. By comparing the data of an early outbreak of a given population with the complete set of historical profiles, the best fit profiles are selected and the corresponding sets of profile codes are used for prediction of the future progression of the epidemic in that population. Application of the method to the data collected for South Korea, Italy and Iran shows that peaks of infection cases are expected to occur before the end of March 2020, and that the percentage of population infected in each city will be less than 0.01%, 0.05% and 0.02%, for South Korea, Italy and Iran, respectively.

## 1. Introduction

The 2019 Coronavirus Disease (COVID-19 or SARS-CoV-2) is a highly contagious disease, which began to spread in China in mid December 2019 [1], and as the volume of intercity travel escalated around the Lunar New Year period, the number of infected individuals began to soar in mid January 2020. With no travel restriction in place due to the low level of vigilance or unawareness of the disease during the early phase of the outbreak, the spreading of the disease had gone almost unobstructed. Travel restriction began to be implemented throughout China since January 24, 2020, which has proven to be effective in curbing the spread of the virus. However, international traffic has not ceased and infectious individuals (who may or may not show any symptom at the time of travel) have actually travelled to different countries with the virus they contracted. Recent studies also show that travel restriction did contribute to the control of the spreading of COVID-19 within China as well as in a global context [2, 3]. By February 20, 2020, China had confirmed a total of 74,579 cases of COVID-19 infection, with death toll reaching 2,119. While China has begun to see declining numbers of infected cases in most cities from late February, other countries started to report surging number of cases in some cities or regions. As of March 6, 2020, the cumulative number of cases of COVID-19 infection was 100,162 worldwide, with South Korea, Italy and Iran reporting surges of infected cases within two weeks. Moreover, the global mortality rate has remained around 3%.

In our recent work (available on February 19, 2020) [4], intercity travel data obtained from Baidu Migration [5] has been collected and integrated into the traditional Susceptible-Exposed-Infected-Removed (SEIR) model [6, 7, 8] to account for the effects of inflow and outflow traffic between 367 cities in China. Parameters of susceptible-to-exposed infection rates, exposed-to-infected infection rates, and recovery rates for 367 cities have been identified by fitting the augmented SEIR model with historical data available from the National Health Commission of China. The predicted spreading profiles of the 367 Chinese cities (which were available on February 19, 2020 [4]) have been highly consistent with the actual profiles, including the times of infection peaks and the percentages of infected individuals in the 367 cities.

In this work, we build on the data and estimated parameters obtained for 367 cities in our prior work [4], and establish a library of profiles (sets of codes) of different spreading profiles. Suppose a new outbreak has occurred in a given population. The numbers of infected and recovered cases during the early phase of the spreading form an initial profile. This initial or incomplete profile is compared with the historical full profiles obtained previously. Then, by selecting the best fit historical profiles, we identify the candidates parameters for prediction of the future spreading profile in that given population (of a city or region). It should be emphasized that the set of historical profiles obtained previously covers various possible spreading dynamics, representing a variety of contact topologies and transmission mechanisms including the travel effects that have been integrated in the model used to capture the transmission dynamics of the 367 cities. Thus, a new outbreak in a given population would likely follow one or a combination of the profiles based on the augmented model in our previous work [4], and hence can be reconstructed from the historical sets of profiles. In this work, we develop a procedure for implementing the selection of historical profiles, identification of best-fit parameters and construction of future profile. We apply the procedure to predict the epidemic progression in the cities of South Korea, Italy and Iran. Results of this study show that the spreading of COVID-19 in most cities in South Korea will peak between March 8 and March 16, 2020, and Italy will peak between March 12 and March 26, 2020, while Iran would have its peak around March 22, 2020. We also investigate the number of infected individuals in each city or region. Our method provides the average number of individuals eventually infected, along with a predicted deviation range at 95% confidence level. For Korea, we predict that Daegu and Gyeongsang North Road would have around 7,619 and 1,287 people eventually infected (i.e., 0.306% and 0.062% of the city’s population), respectively, whereas the number of infected individuals in other cities in South Korea would be fewer than 300, i.e., less than 0.01% of city population. For Italy, we predict that Lombardi and Amelia Romagna would eventually have about 4,784 and 1,555 infected cases (i.e., 0.399% and 0.162 % of region’s population), respectively, and the number of people eventually infected in other cities in Italy would be below 700 (*<*0.05% of city population). Moreover, Iran would have around 14,450 infected individuals, i.e., 0.018% of its population, of which around 2,498 and 1,698 will be expected in Tehran and Zanjan (i.e., 0.39% and 0.2% of the city’s population). In addition, the number of people infected in most other cities would be fewer than 1,000 (*<*0.1% of the city’s population). From the progression trends of the epidemic these three countries, provided control measures continue to be in place, the epidemic would come under control before the end of April 2020.

In the remainder of the paper, we first introduce the official daily infection data used in this study. The augmented SEIR model is briefly reviewed, mainly to introduce the parameters of the model used for prediction of spreading profiles. The key procedure for matching historical profiles and prediction of future spreading profiles will be explained. Results of application of the proposed method to prediction of the peaks and extents of outbreaks in South Korea, Italy and Iran will be given. Finally, we will provide a discussion of our estimation of the propagation and the reasonableness of our estimation in view of the measures taken by the authorities in controlling the spreading of this new disease.

## 2. Data

The World Health Organization currently sets the alert level of COVID-19 to the highest, and has made data related to the epidemic available to the public in a series of situation reports as well as other formats [9]. Our data include the number of infected cases, the cumulative number of infected cases, the number of recovered cases, and death tolls, for individual cities and regions in South Korea, Italy and Mazandran, from February 19, 2020, to March 6, 2020. Data organized in convenient formats are also available elsewhere [10, 11, 12]. Samples of data for Daegu, Gyeongsang North Road (South Korea), Lombardi, Amelia Romagna, Tehran and Iran are shown graphically in Figure 1. It should be noted that the data obtained for South Korea, Italy and Iran correspond to initial stages of the epidemic progression as the number of infected cases are still climbing, as of March 6, 2020.

**Figure 1:**
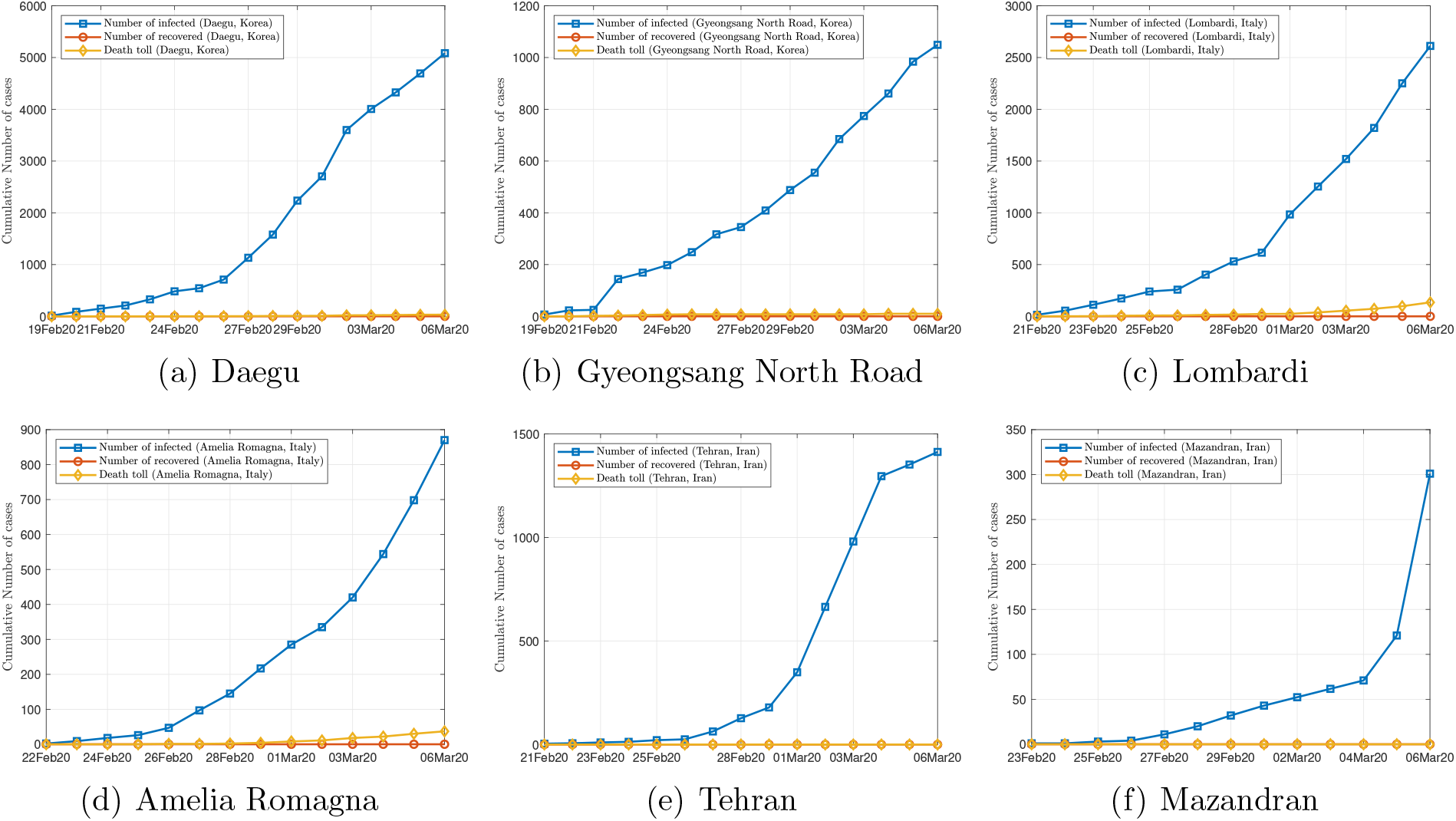
Samples of data.

## 3. Method

### 3.1. The Augmented SEIR Model

The travel-data augmented SEIR model [4] describes the spreading dynamics in terms of a basic fourth-order dynamical system with consideration of intercity travel in China. Consider a city *j* of population *P*_*j*_. The states of the model are the number of susceptible individuals *I*_*j*_(*t*), the number of exposed individuals (infectious but without symptom) *E*_*j*_(*t*), the number of infected individuals *I*_*j*_(*t*), and the number of recovered or removed individuals *R*_*j*_(*t*). The model takes the following form in discrete time [4]:

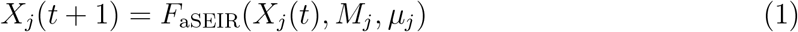

where *X*_*j*_(*t*) = [*S*_*j*_(*t*) *E*_*j*_(*t*) *I*_*j*_(*t*) *R*_*j*_(*t*)]^*T*^ is the state vector on day *t, F*_aSEIR_(.) is the travel-data augmented function, *M*_*j*_ is the set of inflow and outflow travel strengths for city *j*, and *µ*_*j*_ is the set of parameters for city *j*, i.e.,

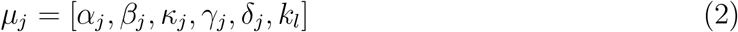

where *β*_*j*_ is the rate at which a susceptible individual is infected by an infected individual in city *j, α*_*j*_ is the rate at which a susceptible individual is infected by an exposed individuals in city *j, κ*_*j*_ is the rate at which an exposed individual becomes infected in city *j*, and *γ*_*j*_ is the recovery rate in city *j, k*_*I*_ is the possibility of an infected individual moving from one city to another, and *δ*_*j*_ is the eventual percentage of the population infected in city *j*. Moreover, the eventual infected population in city *j* is given by 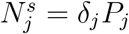.

To facilitate comparison and matching of profiles, we introduce the normalized states as 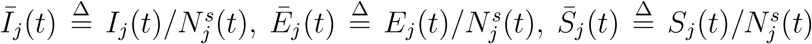 and 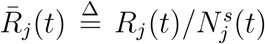. Thus, (1) can be represented in *normalized form* as

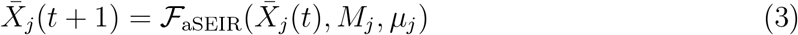

where 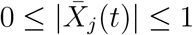. Since the above model has taken into account the human migration effect as well as the necessary transmission mechanism, we may consider the basic set of parameters to represent the characteristics of the propagation profile of city *j*. The complete set of parameters have been identified for 367 cities in China [4], which will serve as a set of codes for various propagation profiles of COVID-19 so far obtained. For brevity, we do not repeat the results here.

While two different cities may have different population size and percentage of eventual infected population, the rates of infection and recovery should be similar across a group of cities, i.e., *µ*_*i*_ *≈µ*_*j*_. Thus, in normalized form, we have

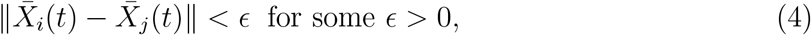

for cities *i* and *j* within a group of cities having similar parameter sets. This also means

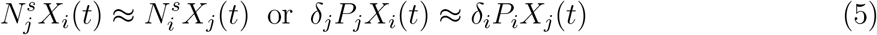

for the group of cities having similar rates of infection and recovery. Thus, provided the historical archive has adequately covered the possible dynamical profiles, we are able to perform fast prediction for any city *o*, by fitting an incomplete set of data (corresponding to an early outbreak stage in city *o*) and using the model parameters already obtained previously, as detailed in the following subsection.

### 3.2. Prediction Method

The proposed data-driven prediction algorithm is based on the set of historical data of the spreading profiles of COVID-19 in 367 cities in China, namely, 367 sets of normalized time series of the form:

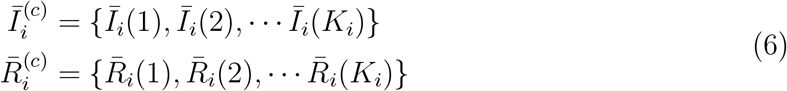

where *i* = 1, 2, *…*, 367, and *K*_*i*_ is the length of the data recorded in city *i*. Superscript “(*c*)” denotes data of Chinese cities.

Now, suppose an outbreak occurs in city *o*, and only *k*_*o*_ days of data have been obtained in normalized form as

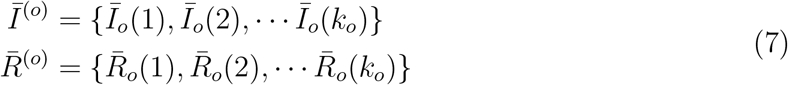

where *k*_*o*_ *< K*_*i*_. Then, assuming the spreading profile of city *o* is related to that of city *i* in the historical archive, as permitted by virtue of the validity of (4), we formulate the following optimization problem to predict the epidemic progression in city *o*:

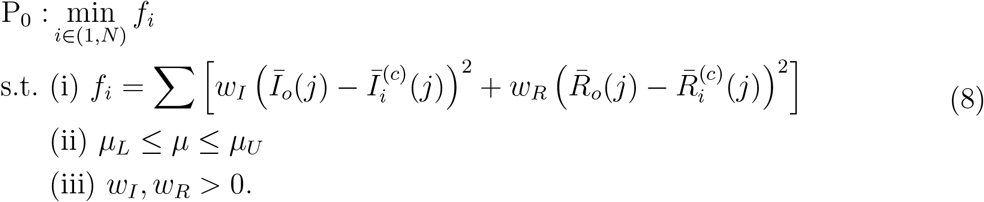

where *N* is the number of Chinese cities in the historical archive, *w*_*I*_ and *w*_*J*_ are weighting coefficients, *µ*_*L*_ and *µ*_*U*_ are the lower and upper bounds of the searching space, respectively. By solving the the nonlinear optimization problem, we can find the most closely resembling growth curve from the historical profiles, e.g., city *i*. Then, we apply the the augmented SEIR model with the profile code given in the parameter set for city *i* to predict the future spreading trend of city *o*. Furthermore, we can choose the top *n* best candidates with the smallest error as the candidate set for prediction, giving an average predicted propagation profile and a deviation range based on *n* best-fit profile codes.

## 4. Results

We apply the aforedescribed procedure to the data obtained so far for cities in South Korea, Italy and Iran. These data represent the early stage of progression of the epidemic spreading, as the trends clearly show that the numbers of infected cases in most cities are still climbing, as of March 6, 2020. For each city or region, we identify a group of 10 profiles of best fit from the historical archive, and retrieve the corresponding sets of profile codes for generating the propagation profiles in the coming days. Using these 10 profiles, we produce an average progression profile, which is also accompanied by a deviation range at 95% confidence level.

Figures 2, 4 and 6 show the data and the predicted number of infected individuals, each with a deviation range of the predicted average trajectory, for South Korea, Italy and Iran, respectively, and Figures 3, 5 and 7 show the corresponding cumulative values. Statistics of infection peaks are shown in Figures 8 (a) to (c). Statistics of the percentage of population eventually infected and the number of individuals eventually infected are shown in Figures 8 (d) to (i). Our key findings are summarized as follows:

1. The number of infected individuals in South Korea, Italy and Iran is expected to continue to increase until mid March 2020. While South Korea will see peaks in most of the cities between March 8, 2020 and March 16, 2020, Italy and Iran are expected to see peaks before the end of March, 2020, as shown in the distributions of the peak times given in Figures 8 (a) to (c).
2. For South Korea, our results show that Daegu and Seoul are the two hardest hit cities, with 7,619 *±*2,096 and 1,287 *±*197 people eventually infected, accounting for 0.306% *±*0.084% and 0.062% *±*0.009% of the city’s population. In other cities in South Korea, the number of infected people will be less than 300, i.e., below 0.01% of the population.
3. For Italy, our results show that Lombardi and Amelia Romagna will have the highest number of cases, with 4,784 *±*788 and 1,555 *±*360 people eventually infected, accounting for 0.399% *±*0.066% and 0.162% *±*0.129% of the region’s population. In other Italian cities or regions, the number of infected people will be below 700, which is less than 0.05% of the population.
4. For Iran, we expect Tehran and Zanjan to be most severely affected, reaching 2,498 *±*566 (0.39% *±*0.02%) and 1,695 *±*92 (0.2% *±*0.07%) cases eventually, respectively. Other Iranian cities will see less than 1,000 eventual infected cases. Our prediction gives a total of 14450 *±*6244 individuals eventually infected, which is 0.018% *±*0.008% of the country’s population.
5. Provided the authorities continue to impose strict control measures, the epidemic will come under control by the end of April and is expected to end before June 2020, and as the quality of treatment improves, more rapid recovery will be expected.

**Figure 2:**
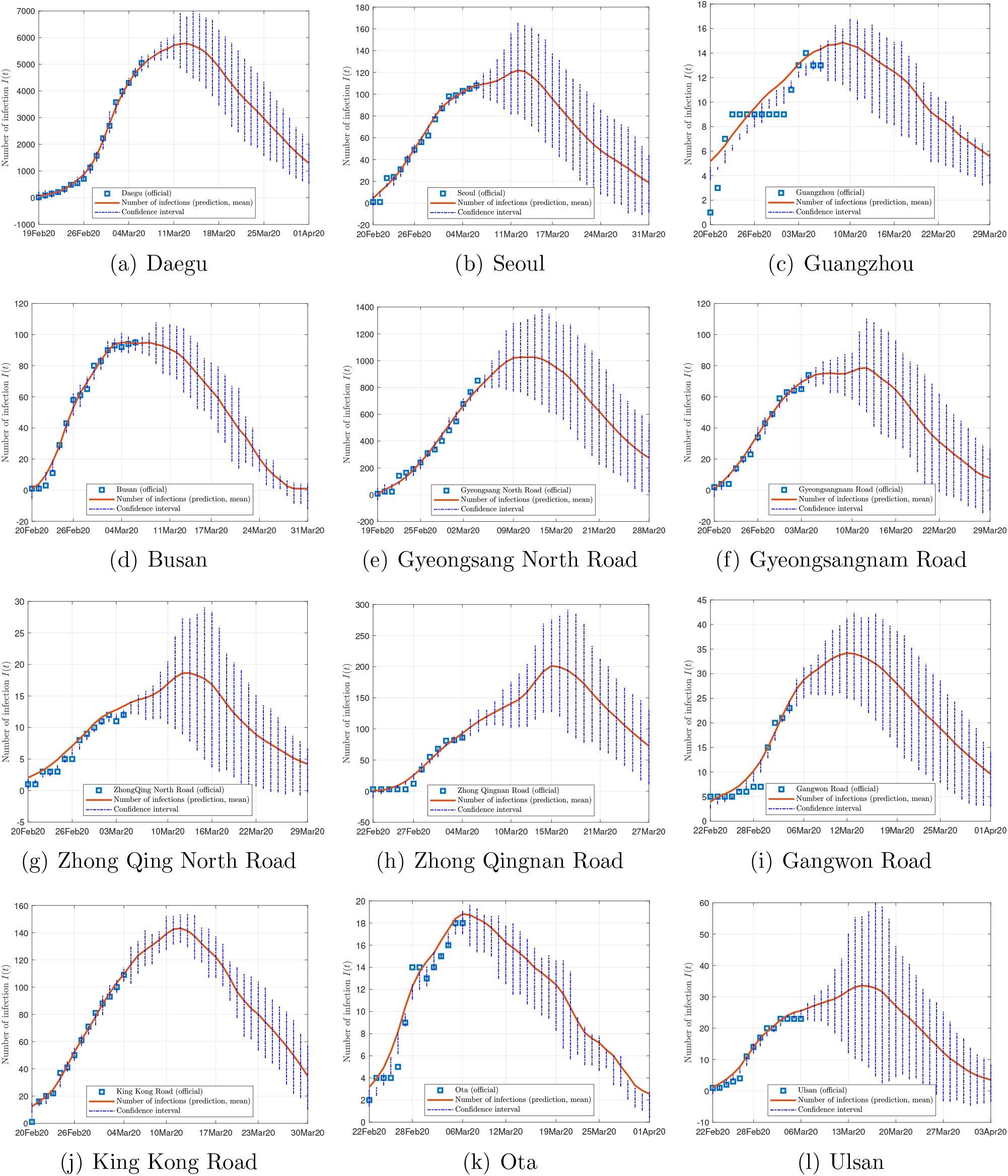
Official and estimated number of infected individuals in cities or regions in South Korea.

**Figure 3:**
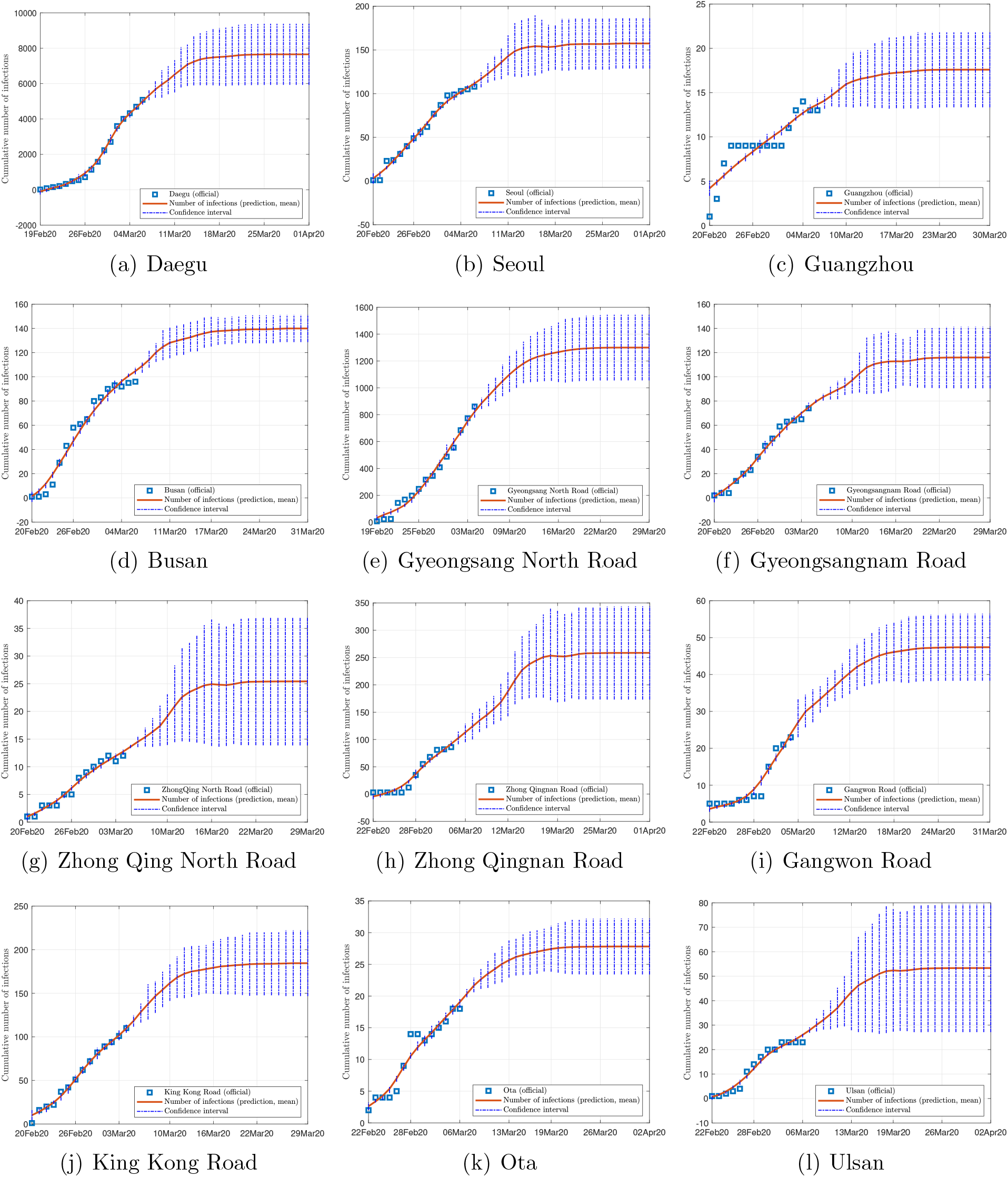
Official and estimated cumulative number of infected individuals in cities or regions in South Korea.

**Figure 4:**
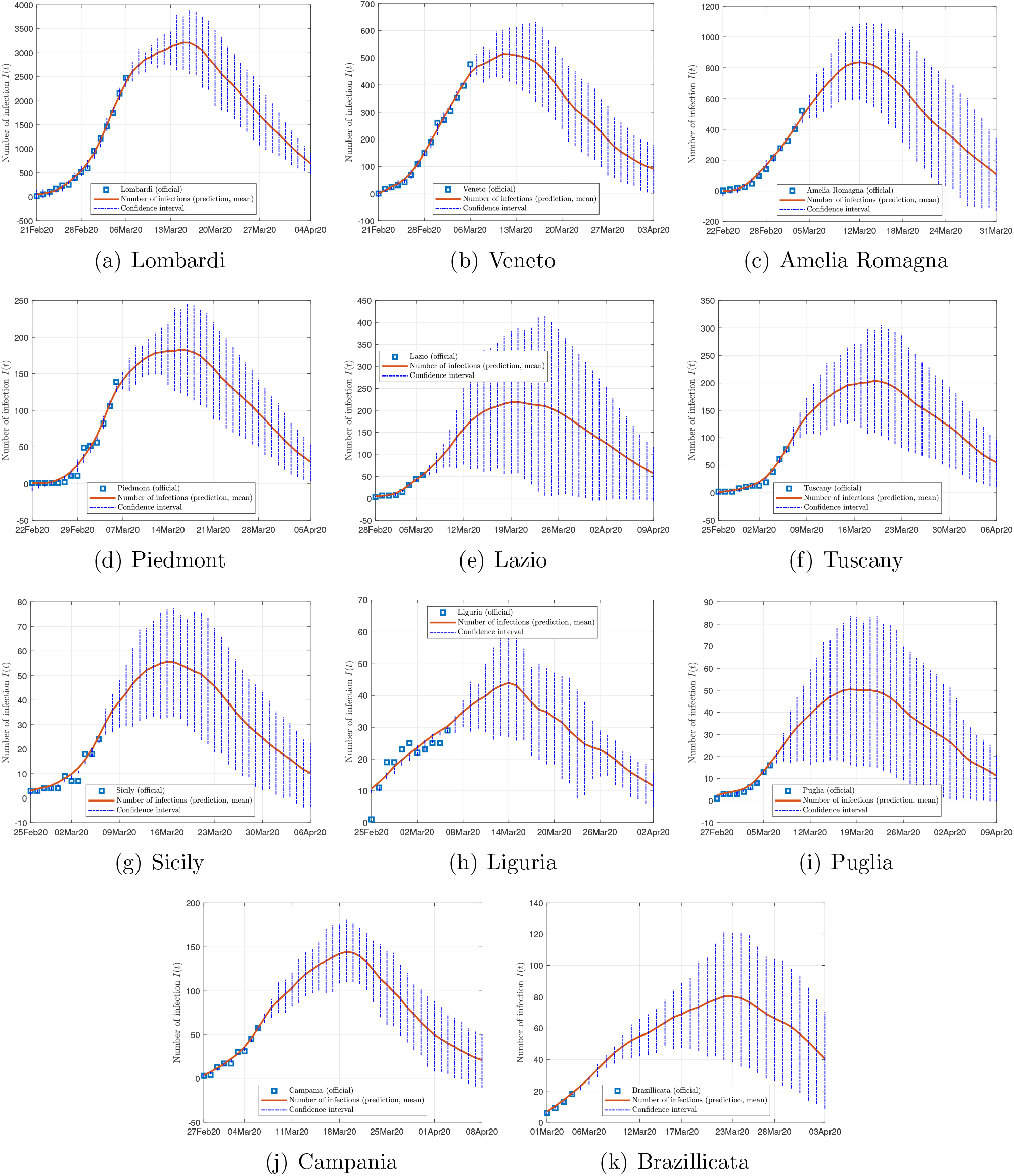
Official and estimated number of infected individuals in regions in Italy.

**Figure 5:**
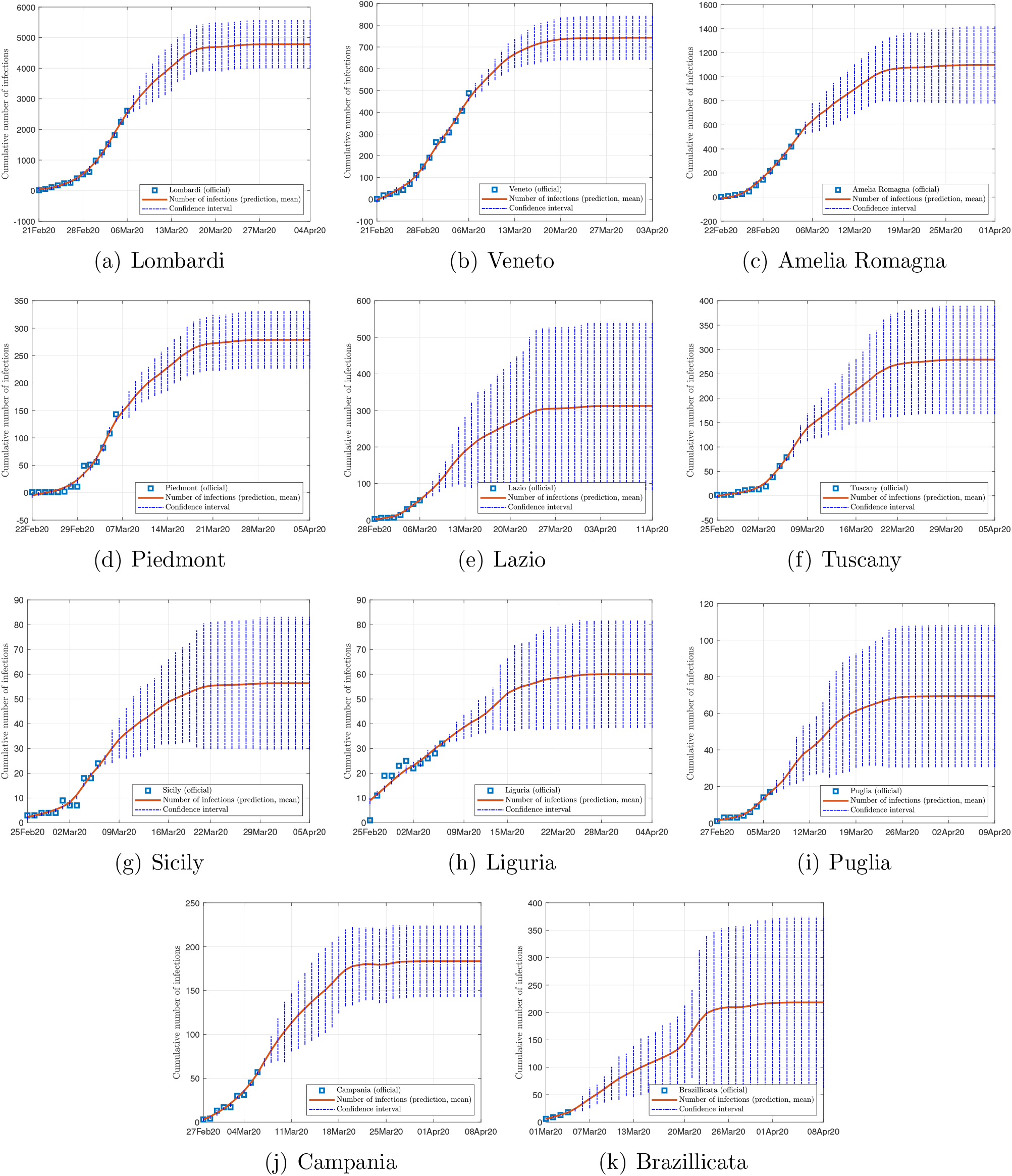
Official and estimated cumulative number of infected individuals in regions in Italy.

**Figure 6:**
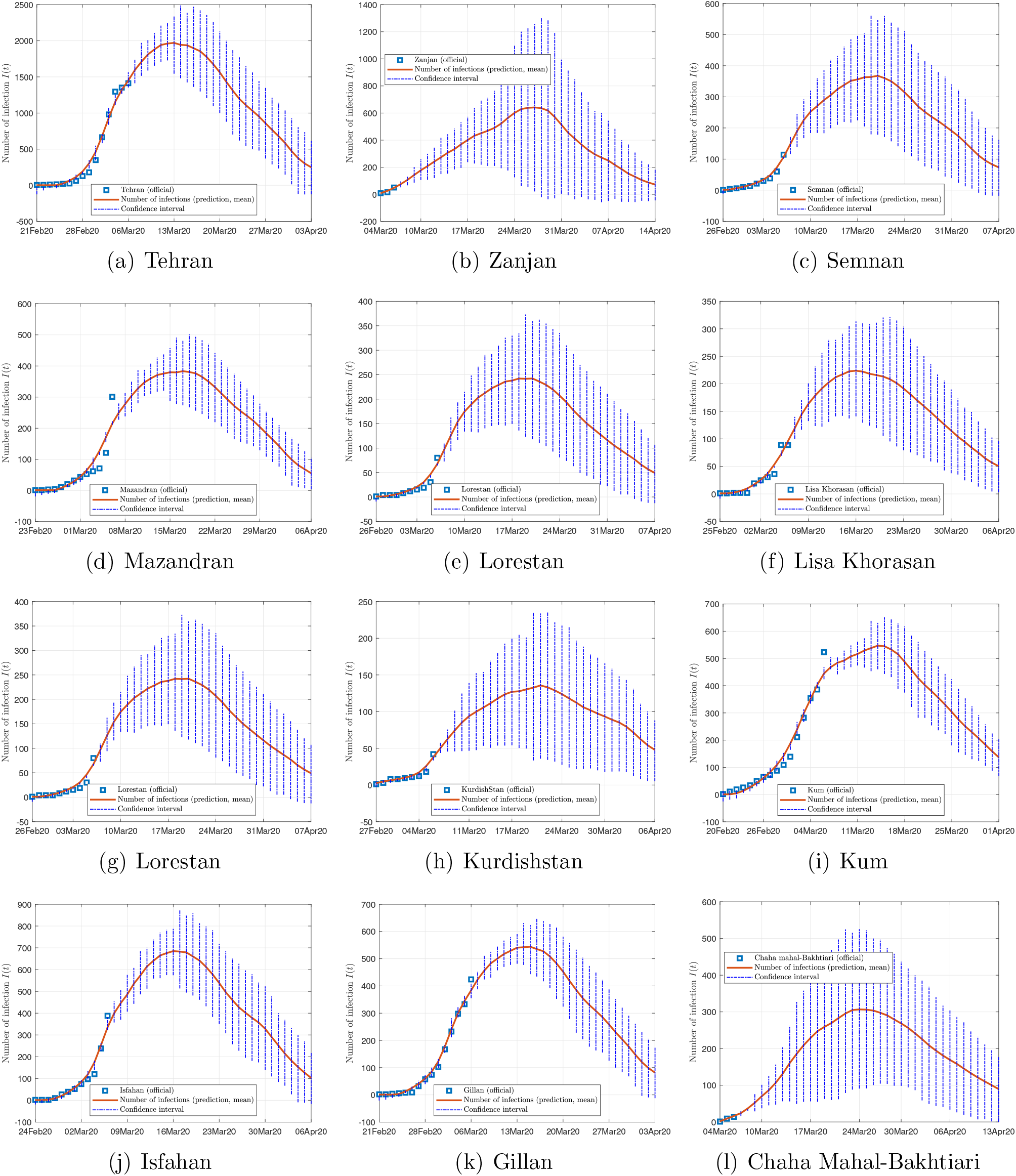
Official and estimated number of infected individuals in some cities in Iran.

**Figure 7:**
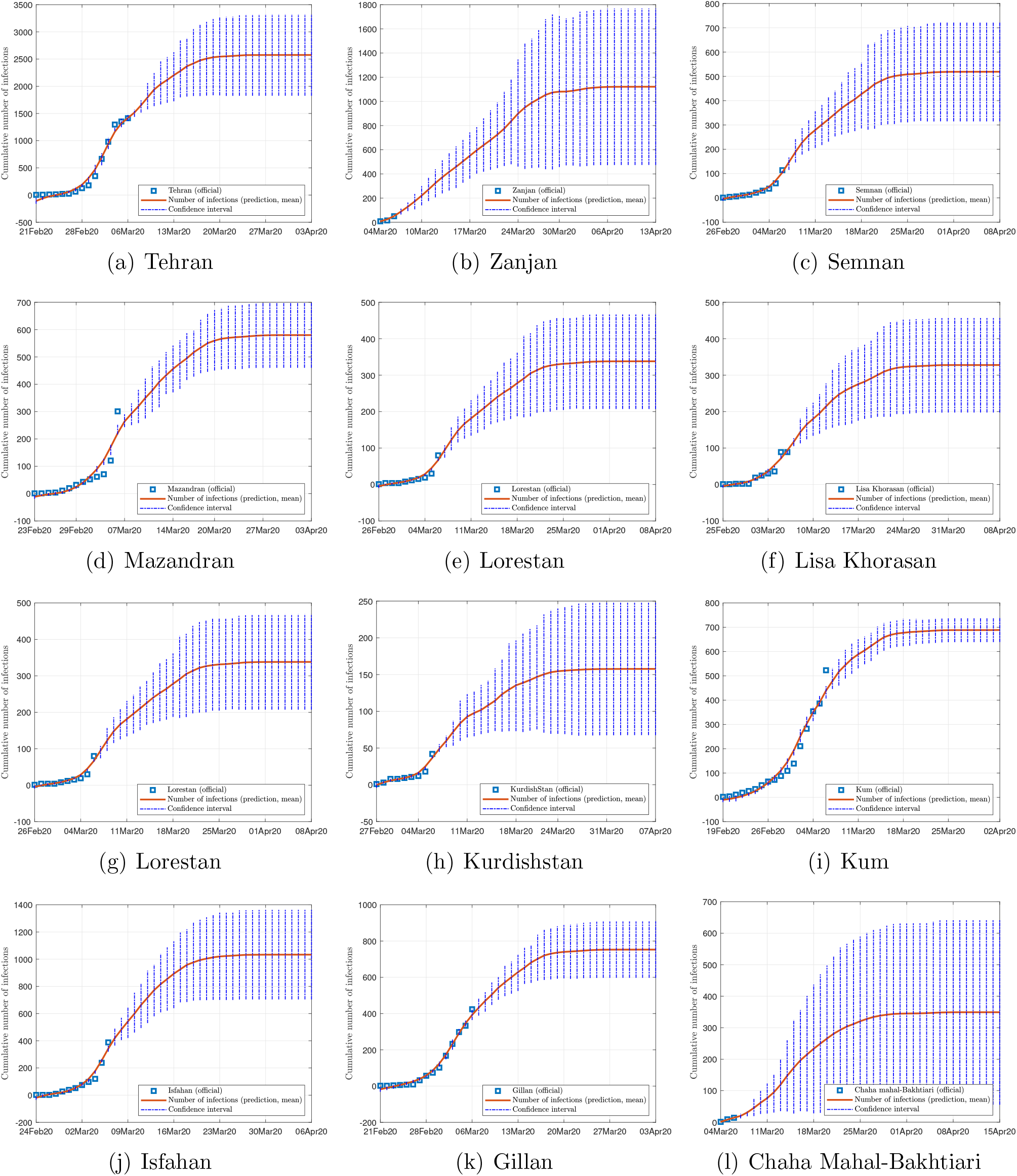
Official and estimated cumulative number of infected individuals in some cities in Iran.

**Figure 8:**
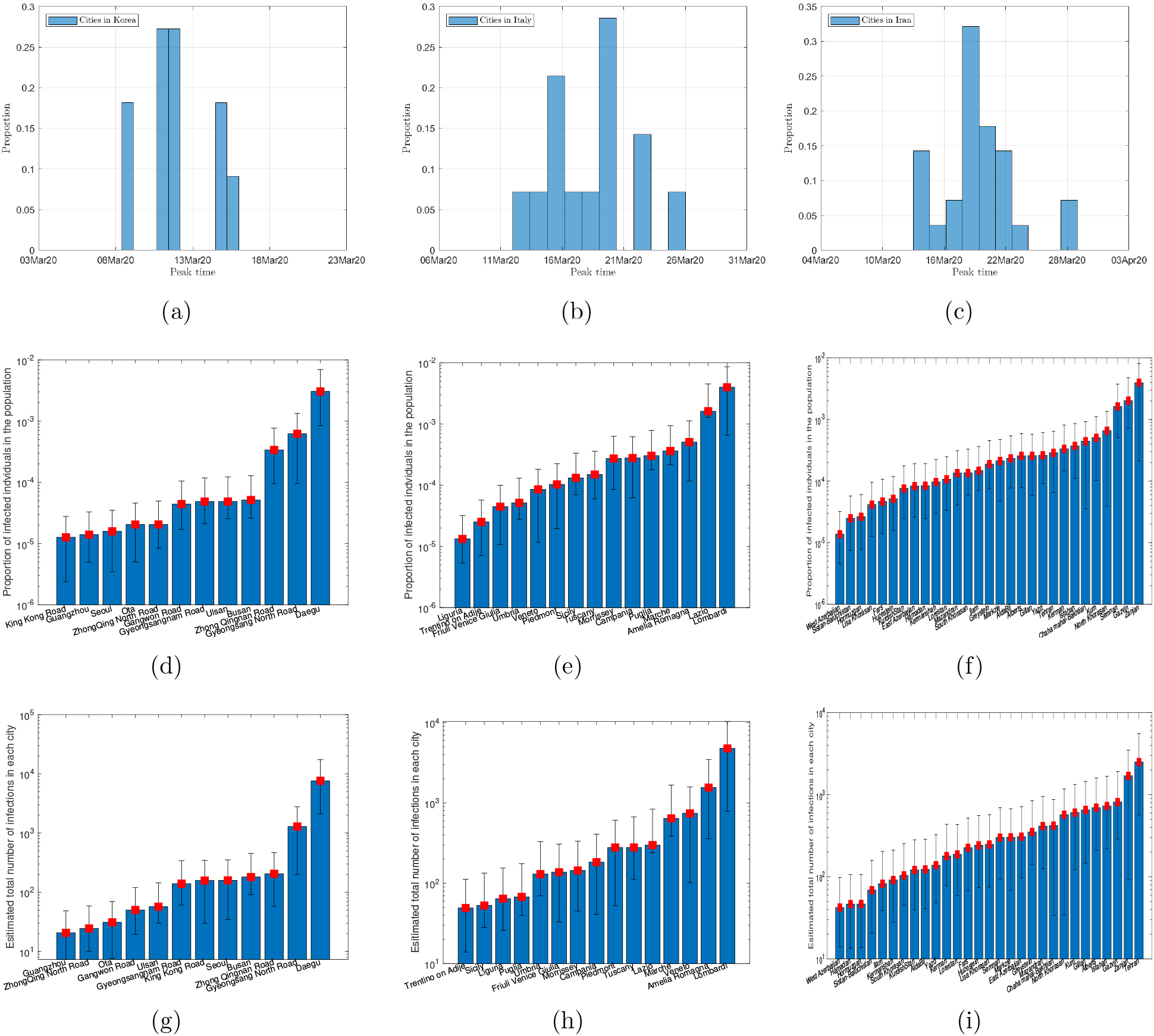
Statistics of (a) peak times in South Korea; (b) peak times in Italy; (c) peak times in Iran; (d) proportion of eventual infected population in South Korea; (e) proportion of eventual infected population in Italy; (f) proportion of eventual infected population in Iran; (g) eventual infected population in South Korea; (h) eventual infected population in Italy; (i) eventual infected population in Iran.

Our prediction on the South Korean cities has revealed a very rapid progression of the epidemic, with 5,000 infections emerged within 10 days and peaks to be expected in most cities or regions within about 2 weeks. The Korean authorities have managed to test an overwhelmingly large number of people (140,000) within a short time, thus preventing a large number of infected and infectious individuals not being quarantined in time [13]. This strategy has an obvious advantage of offering a clear picture of the extents and locations of the infected individuals in the country at the early phase of the epidemic progression. The epidemic progression is found to be more rapid than typical, reflecting on the effectiveness of the control measures being taken.

Italy has the second highest death toll after China, reaching 197 on March 6, 2020 [14]. The fatality rate is about 4%, which is the highest in the world. With infection cases soaring to 3,916 (as of March 6, 2020), Italy had implemented control measures to contain the spread of the virus by shutting down schools and suspending public events in regions where outbreaks were reported. The epidemic is expected to progress in a typical pace (with the present set of parameters), unless more stringent measures are in place.

The situation in Iran is also critical, with the number of infected cases escalated to over 4,000 in less than 2 weeks. Iran has reported death of two lawmakers as of March 7, 2020, and has been struggling to control the contagion, which has spread to 31 provinces [15]. The progression profile is again typical, however, expecting to peak in around 3 weeks. Like Italy, most cities show typical spreading profiles, and the peaks and subsequent decline are not expected to advance sooner unless more stringent measures are implemented to control the contagion.

Finally, depending on the effectiveness of treatment, recovery rates vary, and judging from the predicted trends shown in Figures 2, 4 and 6, the epidemic progressions for the three countries are expected to subside by the end of May, with South Korea expected to recover sooner than the others.

## 5. Conclusion

The spreading of the 2019 New Coronavirus Disease (COVID-19 or SARS-CoV-2) has evolved from a contagion originally confined within Wuhan, China, in December 2019, rapidly to a global contagion, which has spread to 87 countries within two months. The numbers of confirmed infection cases in South Korea, Italy and Iran have surged in the last two weeks, reaching 7,313, 5,883 and 5,823, respectively, on March 8, 2020. The global fatality rate, however, remains below 3%. In this study, we build on the result of our previous work [4] that establishes a library of parameters of an augmented SEIR model, corresponding to the historic spreading profiles of 367 cities in China. This library forms a set of profile codes that cover a variety of possible epidemic progression profiles. By comparing the early incomplete data of epidemic progression collected for a specific population with the historic profiles, we select a few candidate profiles from the historic archive using a nonlinear optimization procedure. The corresponding profile codes of the selected historic progression profiles can then be used to produce estimates of the future progression for that specific population. We apply this method to predict the spreading of COVID-19 in South Korea, Italy and Iran. Results show that the three countries will soon see infection peaks in most cities in the coming 2 to 3 weeks, with South Korea’s cases reaching their peaks slightly earlier than the others. The percentage of population eventually infected will be less than 0.3%, 0.05% and 0.02% for South Korea, Italy and Iran, respectively. The epidemic is expected to end before June 2020, and depending on the effectiveness of treatment, particular cities may see full recovery or zero infection sooner or later than others. It is worth noting that the epidemic progression in South Korean cities are found to be more rapid than typical, implying that the authorities might have taken effective measures to control the spread. The predicted progressions for Italy and Iran, on the other hand, are found to display profiles that are typical of those in the historical archive, and unless more stringent measures are taken, the peaks and subsequent decline of the infection numbers will unlikely come sooner or more rapidly than the predicted trajectories. Finally, we should stress that the proposed data-driven coding method is applicable to predicting epidemic progression in any given population and the accuracy of prediction will depend on the adequacy of the available data in allowing a reliable match to be identified from the historical archive.

## Data Availability

All data referred to in the manuscript will be made available upon request. Updated daily COVID-19 infection data are available from WHO at https://www.who.int/emergencies/diseases/novel-coronavirus-2019

https://en.wikipedia.org/wiki2020\_coronavirus\_outbreak\_in\_South\_Korea

## Acknowledgment

This work was supported by National Science Foundation of China Project 61703355, Guangdong Youth University Innovative Talents Project 2016KQNCX223, and City University of Hong Kong under Special Fund 9380114.

